# Using excess deaths and testing statistics to improve estimates of COVID-19 mortalities

**DOI:** 10.1101/2021.01.10.21249524

**Authors:** Lucas Böttcher, Maria R. D’Orsogna, Tom Chou

## Abstract

Factors such as non-uniform definitions of mortality, uncertainty in disease prevalence, and biased sampling complicate the quantification of fatality during an epidemic. Regardless of the employed fatality measure, the infected population and the number of infection-caused deaths need to be consistently estimated for comparing mortality across regions. We combine historical and current mortality data, a statistical testing model, and an SIR epidemic model, to improve estimation of mortality. We find that the average excess death across the entire US is 13% higher than the number of reported COVID-19 deaths. In some areas, such as New York City, the number of weekly deaths is about eight times higher than in previous years. Other countries such as Peru, Ecuador, Mexico, and Spain exhibit excess deaths significantly higher than their reported COVID-19 deaths. Conversely, we find negligible or negative excess deaths for part and all of 2020 for Denmark, Germany, and Norway.

## Introduction

The novel severe acute respiratory syndrome coronavirus 2 (SARS-CoV-2) first identified in Wuhan, China in December 2019 quickly spread across the globe, leading to the declaration of a pandemic on March 11, 2020 [1]. The emerging disease was termed COVID-19. As of this January 2020 writing, more than 86 million people have been infected, and more than 1.8 million deaths from COVID-19 in more than 218 countries [2] have been confirmed. About 61 million people have recovered globally.

Properly estimating the severity of any infectious disease is crucial for identifying near-future scenarios, and designing intervention strategies. This is especially true for SARS-CoV-2 given the relative ease with which it spreads, due to long incubation periods, asymptomatic carriers, and stealth transmissions [3]. Most measures of severity are derived from the number of deaths, the number of confirmed and unconfirmed infections, and the number of secondary cases generated by a single primary infection, to name a few. Measuring these quantities, determining how they evolve in a population, and how they are to be compared across groups, and over time, is challenging due to many confounding variables and uncertainties.

For example, quantifying COVID-19 deaths across jurisdictions must take into account the existence of different protocols in assigning cause of death, cataloging co-morbidities [4], and lag time reporting [5]. Inconsistencies also arise in the way deaths are recorded, especially when COVID-19 is not the direct cause of death, rather a co-factor leading to complications such as pneumonia and other respiratory ailments [6]. In Italy, the clinician’s best judgment is called upon to classify the cause of death of an untested person who manifests COVID-19 symptoms. In some cases, such persons are given post-mortem tests, and if results are positive, added to the statistics. Criteria vary from region to region [7]. In Germany, postmortem testing is not routinely employed, possibly explaining the large difference in mortality between the two countries. In the US, current guidelines state that if typical symptoms are observed, the patient’s death can be registered as due to COVID-19 even without a positive test [8]. Certain jurisdictions will list dates on which deaths actually occurred, others list dates on which they were reported, leading to potential lag-times. Other countries tally COVID-19 related deaths only if they occur in hospital settings, while others also include those that occur in private and/or nursing homes.

In addition to the difficulty in obtaining accurate and uniform fatality counts, estimating the prevalence of the disease is also a challenging task. Large-scale testing of a population where a fraction of individuals is infected, relies on unbiased sampling, reliable tests, and accurate recording of results. One of the main sources of systematic bias arises from the tested subpopulation: due to shortages in testing resources, or in response to public health guidelines, COVID-19 tests have more often been conducted on symptomatic persons, the elderly, front-line workers and/or those returning from hot-spots. Such non-random testing overestimates the infected fraction of the population.

Different types of tests also probe different infected subpopulations. Tests based on reverse-transcription polymerase chain reaction (RT-PCR), whereby viral genetic material is detected primarily in the upper respiratory tract and amplified, probe individuals who are actively infected. Serological tests (such as enzyme-linked immunosorbent assay, ELISA) detect antiviral antibod-ies and thus measure individuals who have been infected, including those who have recovered.

Finally, different types of tests exhibit significantly different “Type I” (false positive) and “Type II” (false negative) error rates. The accuracy of RT-PCR tests depends on viral load which may be too low to be detected in individuals at the early stages of the infection, and may also depend on which sampling site in the body is chosen. Within serological testing, the kinetics of antibody response are still largely unknown and it is not possible to determine if and for how long a person may be immune from reinfection. Instrumentation errors and sample contamination may also result in a considerable number of false positives and/or false negatives. These errors confound the inference of the infected fraction. Specifically, at low prevalence, Type I false positive errors can significantly bias the estimation of the IFR.

Other quantities that are useful in tracking the dynamics of a pandemic include the number of recovered individuals, tested, or untested. These quantities may not be easily inferred from data and need to be estimated from fitting mathematical models such as SIR-type ODEs [9], age-structured PDEs [10], or network/contact models [11–13].

Administration of tests and estimation of all quantities above can vary widely across jurisdictions, making it difficult to properly compare numbers across them. In this paper, we incorporate excess death data, testing statistics, and mathematical modeling to self-consistently compute and compare mortality across different jurisdictions. In particular, we will use excess mortality statistics [14–16] to infer the number of COVID-19-induced deaths across different regions. We then present a statistical testing model to estimate jurisdiction-specific infected fractions and mortalities, their uncertainty, and their dependence on testing bias and errors. Our statistical analyses and source codes are available at [17].

## Methods

### Mortality measures

Many different fatality rate measures have been defined to quantify epidemic outbreaks [18]. One of the most common is the case fatality ratio (CFR) defined as the ratio between the number of confirmed “infection-caused” deaths *D*_c_ in a specified time window and the number of infections *N*_c_ confirmed within the same time window, CFR = *D*_c_*/N*_c_ [19]. Depending on how deaths *D*_c_ are counted and how infected individuals *N*_c_ are defined, the operational CFR may vary. It may even exceed one, unless all deaths are tested and included in *N*_c_.

Another frequently used measure is the infection fatality ratio (IFR) defined as the true number of “infection-caused” deaths *D* = *D*_c_ + *D*_u_ divided by the actual number of cumulative infections to date, *N*_c_ + *N*_u_. Here, *D*_u_ is the number of unreported infection-caused deaths within a specified period, and *N*_u_ denotes the untested or unreported infections during the same period. Thus, IFR = *D/*(*N*_c_ + *N*_u_).

One major issue of both CFR and IFR is that they do not account for the time delay between infection and resolution. Both measures may be quite inaccurate early in an outbreak when the number of cases grows faster than the number of deaths and recoveries [10]. An alternative measure that avoids case-resolution delays is the confirmed resolved mortality *M* = *D*_c_*/*(*D*_c_ + *R*_c_) [10], where *R*_c_ is the cumulative number of confirmed recovered cases evaluated in the same specified time window over which *D*_c_ is counted. One may also define the true resolved mortality via ℳ = *D/*(*D* + *R*), the proportion of the actual number of deaths relative to the total number of deaths and recovered individuals during a specified time period. If we decompose *R* = *R*_c_ +*R*_u_, where *R*_c_ are the confirmed and *R*_u_, the unreported recovered cases, ℳ= (*D*_c_ + *D*_u_)*/*(*D*_c_ + *D*_u_ + *R*_c_ + *R*_u_). The total confirmed population is defined as *N*_c_ = *D*_c_ + *R*_c_ + *I*_c_, where *I*_c_ the number of living confirmed infecteds. Applying these definitions to any specified time period (typically from the “start” of an epidemic to the date with the most recent case numbers), we observe that CFR ≤ *M* and IFR ≤ ℳ. After the epidemic has long past, when the number of currently infected individuals *I* approach zero, the two fatality ratios and mortality measures converge if the component quantities are defined and measured consistently, lim_*t→∞*_ CFR(*t*) = lim_*t→∞*_ *M* (*t*) and lim_*t→∞*_ IFR(*t*) = lim_*t→∞*_ *ℳ* (*t*) [10].

The mathematical definitions of the four basic mortality measures *Z* = CFR, IFR, *M, ℳ* defined above are given in Table I and fall into two categories, confirmed and total. Confirmed measures (CFR and *M*) rely only on positive test counts, while total measures (IFR and ℳ) rely on projections to estimate the number of infected persons in the total population *N*. Of the measures listed in Table I, the fatality ratio CFR and confirmed resolved mortality *M* do not require estimates of unreported infections, recoveries, and deaths and can be directly derived from the available confirmed counts *D*_c_, *N*_c_, and *R*_c_ [20]. Estimation of IFR and the true resolved mortality ℳ requires the additional knowledge on the unconfirmed quantities *D*_u_, *N*_u_, and *R*_u_. We describe the possible ways to estimate these quantities, along with the associated sources of bias and uncertainty below.

**TABLE I:** Definitions of mortality measures. Quantities with subscript “c” and “u” denote confirmed (*i*.*e*., positively tested) and unconfirmed populations. For instance, *D*_c_, *R*_c_, and *N*_c_ denote the total number of confirmed dead, recovered, and infected individuals, respectively. *d*^(*j*)^(*i*) is the number of individuals who have died in the *i*^th^ time window (*e*.*g*., day, week) of the *j*^th^ previous year. The mean number of excess deaths between the periods *k*_s_ and *k* this year 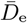 is thus 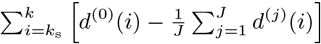. Where the total number of infection-caused deaths *D*_c_ + *D*_u_ appears,it can be estimated using the excess deaths 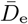 over as detailed in the main text. We have also included raw death numbers/100,000 and the mean excess deaths *r* relative to the mean number of deaths over the same period of time from past years (see Eqs. (1)).

| Measure $Z$<br>Subpopulation | Fatality Ratios | Resolved Mortality | Excess Death Indices |
| --- | --- | --- | --- |
| Confirmed | $\text{CFR} = \frac{D_c}{N_c}$ | $M = \frac{D_c}{D_c + R_c}$ | $D_e \text{ per } 100,000: \frac{D_c + D_u}{100,000}$ |
| Total | $\text{IFR} = \frac{D_c + D_u}{N_c + N_u}$ | $\mathcal{M} = \frac{D_c + D_u}{D_c + D_u + R_c + R_u}$ | relative: $r = \frac{\sum_i \left[ d^{(0)}(i) - \frac{1}{J} \sum_j^J d^{(j)}(i) \right]}{\frac{1}{J} \sum_j^J \sum_i d^{(j)}(i)}$ |

### Excess deaths data

An unbiased way to estimate *D* = *D*_c_ + *D*_u_, the cumulative number of deaths, is to compare total deaths within a time window in the current year to those in the same time window of previous years, before the pandemic. If the epidemic is widespread and has appreciable fatality, one may reasonably expect that the excess deaths can be attributed to the pandemic [21–25]. Within each affected region, these “excess” deaths *D*_e_ relative to “historical” deaths, are independent of testing limitations and do not suffer from highly variable definitions of virus-induced death. Thus, within the context of the COVID-19 pandemic, *D*_e_ is a more inclusive measure of virus-induced deaths than *D*_c_ and can be used to estimate the total number of deaths, 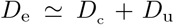. Moreover, using data from multiple past years, one can also estimate the uncertainty in *D*_e_. In practice, deaths are typically tallied daily, weekly [21, 28], or sometimes aggregated monthly [27, 29] with historical records dating back *J* years so that for every period *i* there are a total of *J* + 1 death values. We denote by *d*^(*j*)^(*i*) the total number of deaths recorded in period *i* from the *j*^th^ previous year where 0 ≤ *j* ≤ *J* and where *j* = 0 indicates the current year. In this notation, 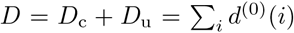, where the summation tallies deaths over several periods of interest within the pandemic. Note that we can decompose 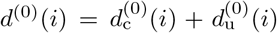, to include the contribution from the confirmed and unconfirmed deaths during each period *i*, respectively. To quantify the total cumulative excess deaths we derive excess deaths 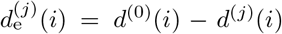 per week relative to the *j*^th^ previous year. Since *d*^(0)^(*i*) is the total number of deaths in week *i* of the current year, by definition 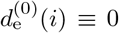. The excess deaths during week *i*, 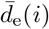, averaged over *J* past years and the associated, unbiased variance *σ*_e_(*i*) are given by

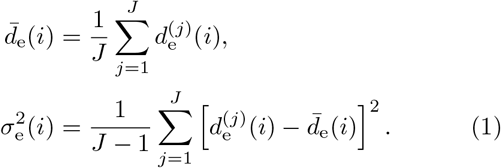

The corresponding quantities accumulated over *k* weeks define the mean and variance of the cumulative excess deaths 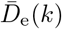 and Σ_e_(*k*)

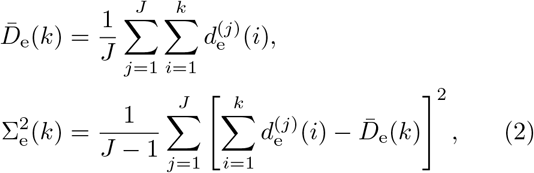

where deaths are accumulated from the first to the *k*^th^ week of the pandemic. The variance in Eqs. (1) and (2) arise from the variability in the baseline number of deaths from the same time period in *J* previous years.

We gathered excess death statistics from over 23 countries and all US states. Some of the data derive from open-source online repositories as listed by official statistical bureaus and health ministries [21–25, 29]; other data are elaborated and tabulated in Ref. [27]. In some countries excess death statistics are available only for a limited number of states or jurisdictions (*e*.*g*., Brazil). The US death statistics that we use in this study is based on weekly death data between 2015–2019 [29]. For all other countries, the data collection periods are summarized in Ref. [27]. Fig. A1(a-b) shows historical death data for NYC and Germany, while Fig. A1(c-d) plots the confirmed and excess deaths and their confidence levels computed from Eqs. (1) and (2). We assumed that the cumulative summation is performed from the start of 2020 to the current week *k* = *K* so that 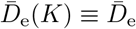 indicates excess deaths at the time of writing. Significant numbers of excess deaths are clearly evident for NYC, while Germany thus far has not experienced significant excess deaths.

**Fig. 1:**
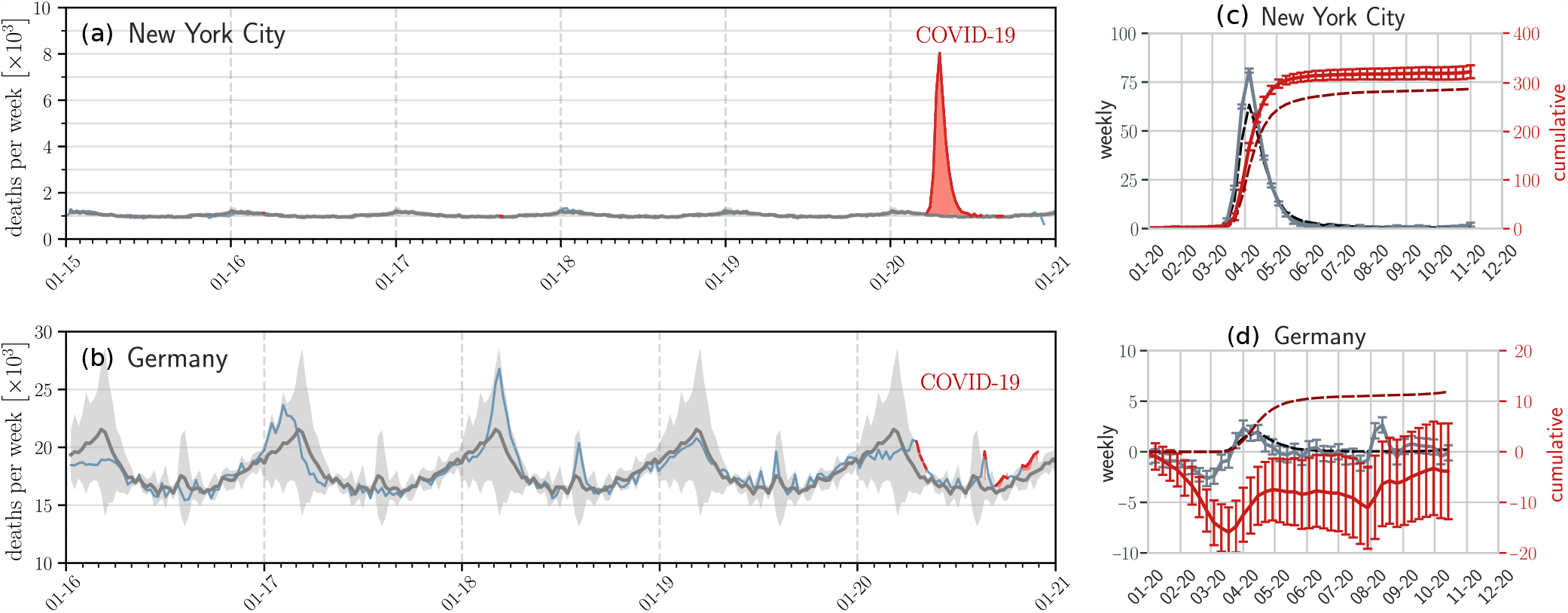
Examples of seasonal mortality and excess deaths. The evolution of weekly deaths in (a) New York City (six years) and (b) Germany (five years) derived from data in Refs. [26, 27]. Grey solid lines and shaded regions represent the historical numbers of deaths and corresponding confidence intervals defined in Eq. (1). Blue solid lines indicate weekly deaths, and weekly deaths that lie outside the confidence intervals are indicated by solid red lines. The red shaded regions represent statistically significant mean cumulative excess deaths *D*_e_. The reported weekly confirmed deaths 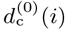 (dashed black curves), reported cumulative confirmed deaths *D*_c_(*k*) (dashed dark red curves), weekly excess deaths 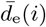 (solid grey curves), and cumulative excess deaths 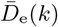 (solid red curves) are plotted in units of per 100,000 in (c) and (d) for NYC and Germany, respectively. The excess deaths and the associated 95% confidence intervals given by the error bars are constructed from historical death data in (a-b) and defined in Eqs. (1) and (2). In NYC there is clearly a significant number of excess deaths that can be safely attributed to COVID-19, while to date in Germany, there have been no significant excess deaths. Excess death data from other jurisdictions are shown in the Supplementary Information and typically show excess deaths greater than reported confirmed deaths (with Germany an exception as shown in (d)).

To evaluate CFR and *M*, data on only *D*_c_, *N*_c_, and *R*_c_ are required, which are are tabulated by many jurisdictions. To estimate the numerators of IFR and ℳ, we approximate 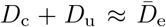 using Eq. (2). For the denominators, estimates of the unconfirmed infected *N*_u_ and unconfirmed recovered populations *R*_u_ are required. In the next two sections we propose methods to estimate *N*_u_ using a statistical testing model and *R*_u_ using compartmental population model.

### Statistical testing model with bias and testing errors

The total number of confirmed and unconfirmed infected individuals *N*_c_ + *N*_u_ appears in the denominator of the IFR. To better estimate the infected population we present a statistical model for testing in the presence of bias in administration and testing errors. Although *N*_c_ + *N*_u_ used to estimate the IFR includes those who have died, depending on the type of test, it may or may not include those who have recovered. If *S, I, R, D* are the numbers of susceptible, currently infected, recovered, and deceased individuals, the total population is *N* = *S* + *I* + *R* + *D* and the infected fraction can be defined as *f* = (*N*_c_ + *N*_u_)*/N* = (*I* + *R* + *D*)*/N* for tests that include recovered and deceased individuals (*e*.*g*., antibody tests), or *f* = (*N*_c_ + *N*_u_)*/N* = (*I* + *D*)*/N* for tests that only count currently infected individuals (*e*.*g*., RTPCR tests). If we assume that the total population *N* can be inferred from census estimates, the problem of identifying the number of unconfirmed infected persons *N*_u_ is mapped onto the problem of identifying the true fraction *f* of the population that has been infected.

Typically, *f* is determined by testing a representative sample and measuring the proportion of infected persons within the sample. Besides the statistics of sampling, two main sources of systematic errors arise: the non-random selection of individuals to be tested and errors intrinsic to the tests themselves. Biased sampling arises when testing policies focus on symptomatic or at-risk individuals, leading to over-representation of infected individuals.

Figure 2 shows a schematic of a hypothetical initial total population of *N* = 54 individuals in a specified jurisdiction. Without loss of generality we assume there are no unconfirmed deaths, *D*_u_ = 0, and that all confirmed deaths are equivalent to excess deaths, so that 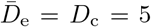 in the jurisdiction represented by Fig. 2. Apart from the number of deceased, we also show the number of infected and uninfected subpopulations and label them as true positives, false positives, and false negatives. The true number of infected individuals is *N*_c_ + *N*_u_ = 16 which yields the true *f* = 16*/*54 = 0.27 and an IFR = 5/16 = 0.312 within the jurisdiction.

**Fig. 2:**
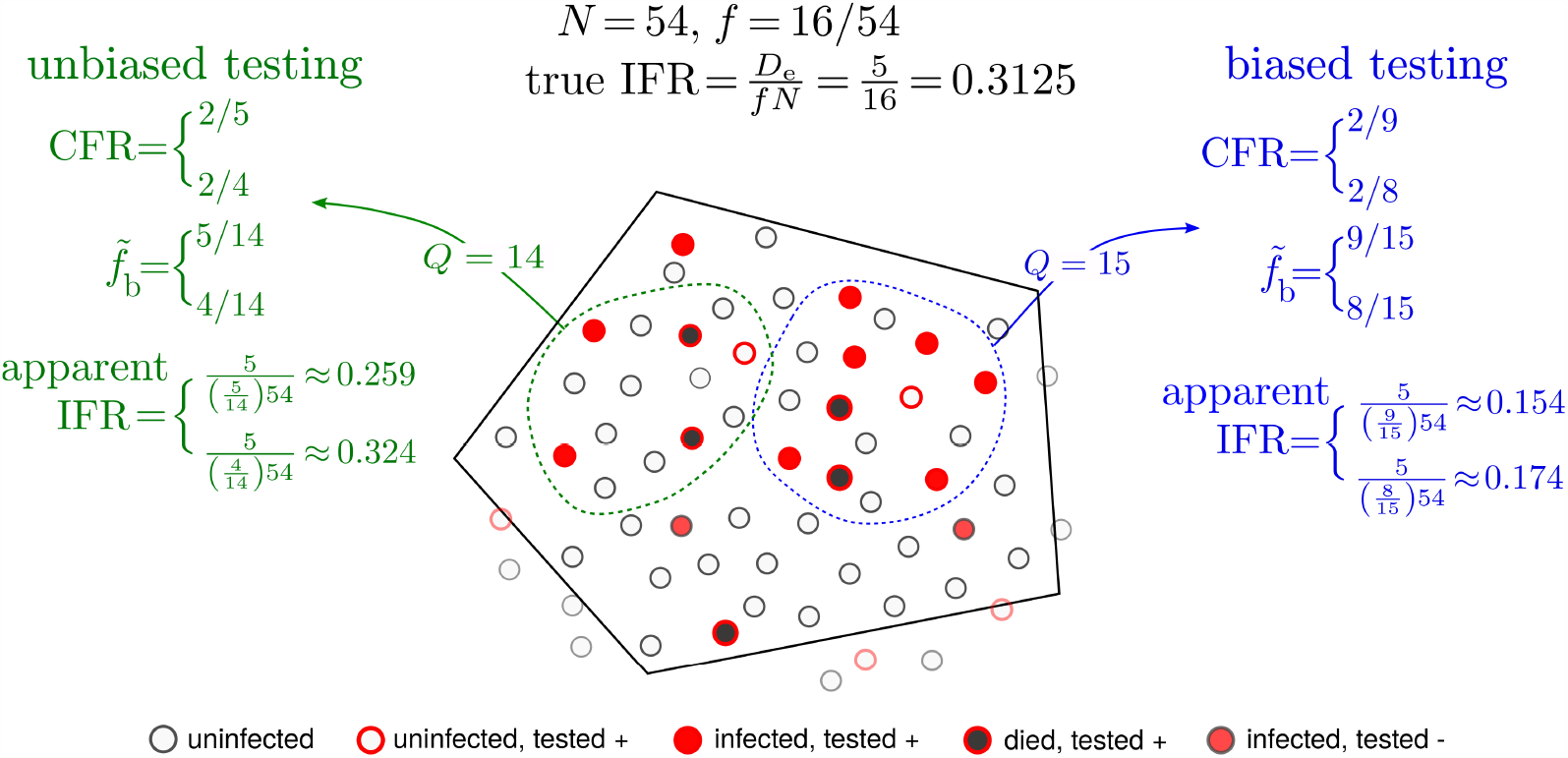
Biased and unbiased testing of a population. A hypothetical scenario of testing a population (total *N* = 54 individuals) within a jurisdiction (solid black boundary). Filled red circles represent the true number of infected individuals who tested positive and the black-filled red circles indicate individuals who have died from the infection. Open red circles denote uninfected individuals who were tested positive (false positives) while filled red circles with dark gray borders are infected individuals who were tested negative (false negatives). In the jurisdiction of interest 5 have died of the infection while 16 are truly infected. The true fraction *f* of infected in the entire population is thus *f* = 16*/*54 and the true IFR=5*/*16. However, under testing (green and blue) samples, a false positive is shown to arise. If the apparent positive fraction 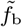 is derived from a biased sample (blue), the estimated *apparent* IFR can be quite different from the true one. For a less biased (more random) testing sample (green sample), a more accurate estimate of the total number of infected individuals is 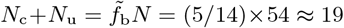 when the single false positive in this sample is included, and 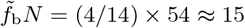 when the false positive is excluded, and allows us to more accurately infer the IFR. Note that CFR is defined according to the tested quantities *D*_c_*/N*_c_ which are precisely 2*/*9 and 2*/*5 for the blue and green sample, respectively, if false positives are considered. When false negatives are known and factored out CFR = 2*/*8 and 2/4, for the blue and green samples, respectively.

Also shown in Fig. 2 are two examples of sampling. Biased sampling and testing is depicted by the blue contour in which 6 of the 15 are alive and infected, 2 are deceased, and the remaining 7 are healthy. For simplicity, we start by assuming no testing errors. This measured infected fraction of this sample 8*/*15 = 0.533 *> f* = 0.296 is biased since it includes a higher proportion of infected persons, both alive and deceased, than that of the entire jurisdiction. Using this biased measured infected fraction of 8*/*15 yields IFR = 5*/*(0.533 · 54) ≈ 0.174, which significantly underestimates the true IFR = 0.312. A relatively unbiased sample, shown by the green contour, yields an infected fraction of 4*/*14 ≈ 0.286 and an apparent IFR ≈ 0.324 which are much closer to the true fraction *f* and IFR. In both samples discussed above we neglected testing errors such as false positives indicated in Fig. 2. Tests that are unable to distinguish false positives as negatives would yield a larger *N*_c_, resulting in an apparent infected fraction 9*/*15 and an even smaller apparent IFR ≈ 0.154. By contrast, the false positive testing errors on the green sample would yield an apparent infected fraction 5*/*15 = 0.333 and IFR= 0.259.

Given that test administration can be biased, we propose a parametric form for the apparent or measured infected fraction

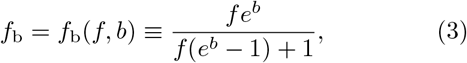

to connect the apparent (biased sampling) infected fraction *f*_b_ with the true underlying infection fraction. The bias parameter −∞*< b <* ∞ describes how an infected or uninfected individual might be preferentially selected for testing, with *b <* 0 (and *f*_b_ *< f*) indicating undertesting of infected individuals, and *b >* 0 (and *f*_b_ *> f*) representing over-testing of infecteds. A truly random, unbiased sampling arises only when *b* = 0 where *f*_b_ = *f*. Given *Q* (possibly biased) tests to date, testing errors, and ground-truth infected fraction *f*, we derive in the SI the likelihood of observing a positive fraction 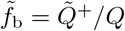 (where 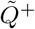 is the number of recorded positive tests):

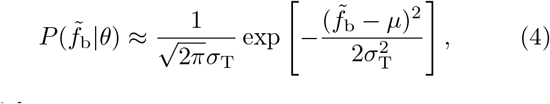

in which

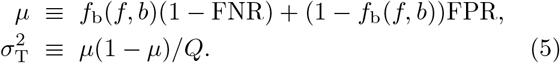

Here, *µ* is the expected value of the measured and biased fraction 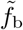 and 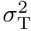 is its variance. Note that the param-eters *θ* = {*Q, f, b*, FPR, FNR} may be time-dependent and change from sample to sample. Along with the likelihood function 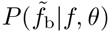, one can also propose a prior distribution *P* (*θ* |*α*) with hyperparameters *α*, and apply Bayesian methods to infer *θ* (see SI).

To evaluate IFR, we must now estimate *f* given 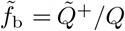 and possible values for FPR, FNR, and/or *b*, or the hyperparameters *α* defining their uncertainty. The simplest maximum likelihood estimate of *f* can be found by maximizing 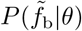 with respect to *f* given a measured value 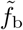 and all other parameter values *θ* specified:

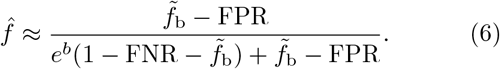

Note that although FNRs are typically larger than FPRs, small values of *f* and 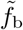 imply that 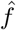 and *µ* are more sensitive to the FPR, as indicated by Eqs. (5) and (6).

If time series data for 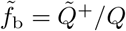 are available, one can evaluate the corrected testing fractions in Eq. (6) for each time interval. Assuming that serological tests can identify infected individuals long after symptom onset, the latest value of 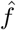 would suffice to estimate corresponding mortality metrics such as the IFR. For RT-PCR testing, one generally needs to track how 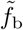 evolves in time. A rough estimate would be to use the mean of 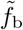 over the whole pandemic period to provide a lower bound of the estimated prevalence 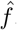.

The measured 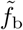 yields only the apparent 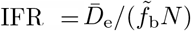, but Eq. (6) can then be used to evaluate the corrected 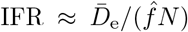 which will be a better estimate of the true IFR. For example, under moderate bias |*b*| ≲ 1 and assuming FNR, FPR, 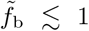 Eq. (6) relates the apparent and corrected IFRs through 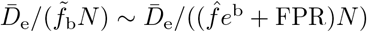.

Another commonly used representation of the IFR is 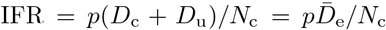. This expression is equivalent to our 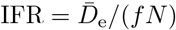 if 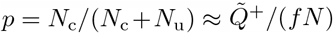 is defined as the fraction of infected individuals that are confirmed [30, 31]. In this alternative representation, the *p* factor implicitly contains the effects of biased testing. Our approach allows the true infected fraction *f* to be directly estimated from 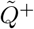 and *N*.

While the estimate 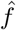 depends strongly on *b* and FPR, and weakly on FNR, the *uncertainty* in *f* will depend on the uncertainty in the values of *b*, FPR, and FNR. A Bayesian framework is presented in the SI, but under a a Gaussian approximation for all distributions, the uncertainty in the testing parameters can be propagated to the squared coeffcient 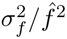 of variation of the estimated infected fraction 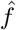, as explicitly computed in the SI. Moreover, the uncertainties in the mortality indices *Z* decomposed into the uncertainties of their individual components are listed in Table II.

**TABLE II:**
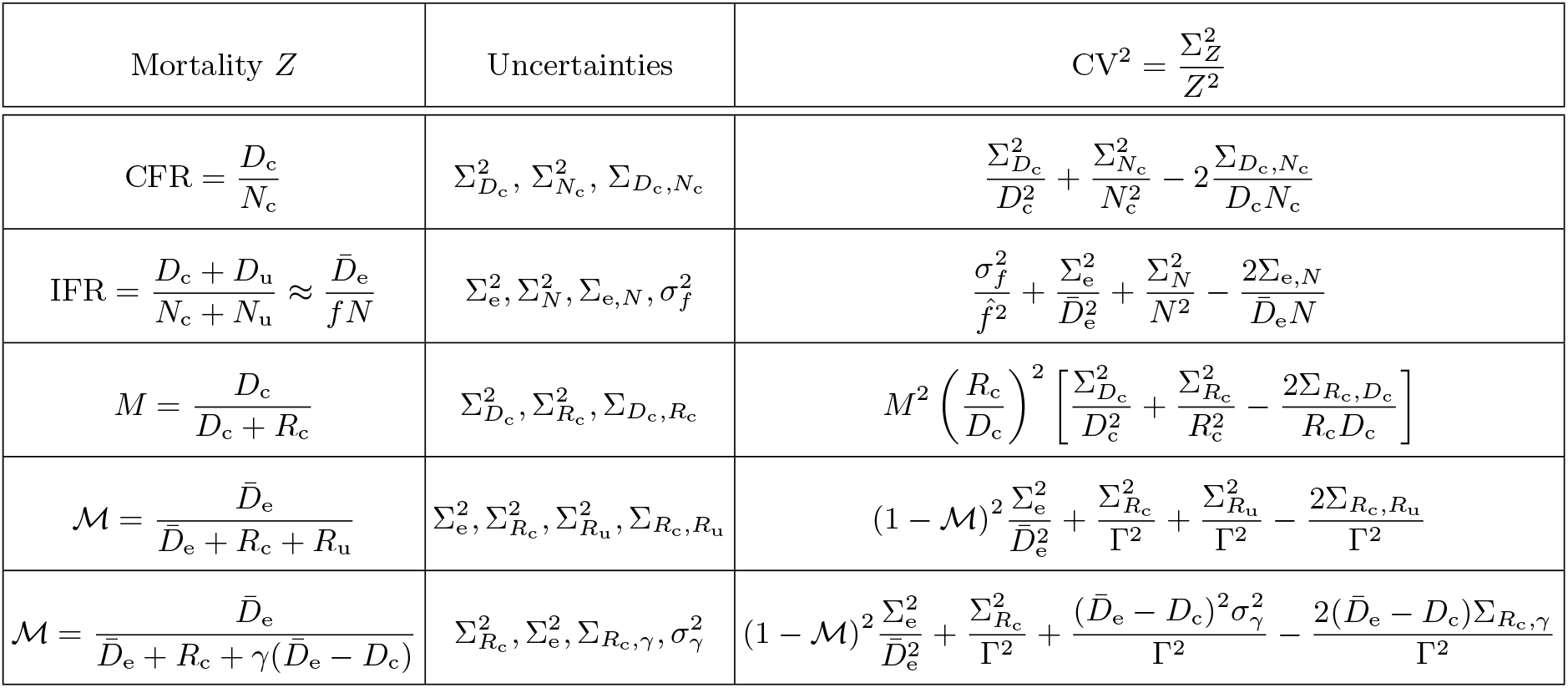
Uncertainty propagation for different mortality measures. Table of squared coefficients of variation 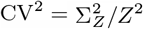 for the different mortality indices *Z* derived using standard error propagation expansions [37]. We use 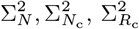, and 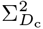 to denote the uncertainties in the total population, confirmed cases, recoveries, and deaths, respectively. The variance of the number of excess deaths is 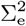, which feature in the IFR and ℳ. The uncertainty in the infected fraction 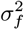 that contributes to the uncertainty in IFR depends on uncertainties in testing bias and testing errors as shown in Eq. (A6). The term 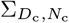 represents the covariance between *D*_c_, *N*_c_, and similarly for all other covariances 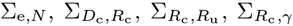. Since variations in *D*_e_ arise from fluctuations in past-year baselines and not from current intrinsic uncertainty, we can neglect correlations between variations in *D*_e_ and uncertainty in *R*_c_, *R*_u_. In the last two rows, representing ℳ expressed in two different ways, 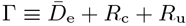 and 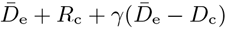, respectively. Moreover, when using the SIR model to replace *D*_u_ and *R*_u_ with 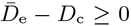, there is no uncertainty associated with *D*_u_ and *R*_u_ in a deterministic model. Thus, covariances cannot be defined except through the uncertainty in the parameter *γ* = *γ*_u_*/µ*_u_.

### Using compartmental models to estimate resolved mortalities

Since the number of unreported recovered individuals *R*_u_ required to calculate ℳ is not directly related to excess deaths nor to positive-tested populations, we use an SIR-type compartmental model to relate *R*_u_ to other inferable quantities [9]. Both unconfirmed recovered individuals and unconfirmed deaths are related to unconfirmed infected individuals who recover at rate *γ*_u_ and die at rate *µ*_u_. The equations for the cumulative numbers of unconfirmed recovered individuals and unconfirmed deaths,

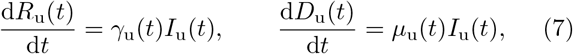

can be directly integrated to find 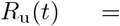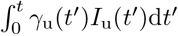 and 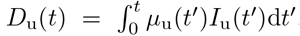 The rates *γ*_u_ and *µ*_u_ may differ from those averaged over the entire population since testing may be biased towards subpopulations with different values of *γ*_u_ and *µ*_u_. If one assumes *γ*_u_ and *µ*_u_ are approximately constant over the period of interest, we find *R*_u_*/D*_u_ ≈ *γ*_u_*/µ*_u_ ≡ *γ*. We now use 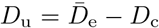, where both 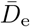 and *D*_c_ are given by data, to estimate 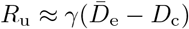 and write ℳ as

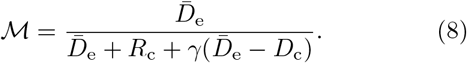

Thus, a simple SIR model transforms the problem of determining the number of unreported death and recovered cases in ℳ to the problem of identifying the recovery and death rates in the untested population. Alterna-tively, we an make use of the fact that both the IFR and resolved mortality ℳ should have comparable values and match ℳ to IFR ≈ 0.1 −1.5% [31–33] by setting *γ* ≡ *γ*_u_*/µ*_u_ ≈ 100 −1000 (see SI for further information). Note that inaccuracies in confirming deaths may give rise to 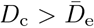. Since by definition, infection-caused excess deaths must be greater than the confirmed deaths, we set 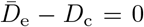 whenever data happens to indicate 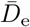 to be less than *D*_c_.

## Results

Here, we present much of the available worldwide fatality data, construct the excess death statistics, and compute mortalities and compare them across jurisdictions. We show that standard mortality measures significantly underestimate the death toll of COVID-19 for most regions (see Figs. A1 and A2). We also use the data to estimate uncertainties in the mortality measures and relate them uncertainties of the underlying components and model parameters.

### Excess and confirmed deaths

We find that in New York City for example, the number of confirmed COVID-19 deaths between March 10, 2020 and December 10, 2020 is 19,694 [34] and thus significantly lower than the 27,938 (95% CI 26,516–29,360) reported excess mortality cases [21]. From March 25, 2020 until December 10, 2020, Spain counts 65,673 (99% confidence interval [CI] 91,816–37,061) excess deaths [22], a number that is substantially larger than the officially reported 47,019 COVID-19 deaths [35]. The large difference between excess deaths and reported COVID-19 deaths in Spain and New York City is also observed in Lombardia, one of the most affected regions in Italy. From February 23, 2020 until April 4, 2020, Lombardia reported 8,656 reported COVID-19 deaths [35] but 13,003 (95% 12,335–13,673) excess deaths [25]. Starting April 5 2020, mortality data in Lombardia stopped being reported in a weekly format. In England/Wales, the number of excess deaths from the onset of the COVID-19 outbreak on March 1, 2020 until November 27, 2020 is 70,563 (95% CI 52,250–88,877) whereas the number of reported COVID-19 deaths in the same time interval is 66,197 [26]. In Switzerland, the number of excess deaths from March 1, 2020 until November 29, 2020 is 5,664 (95% CI 4,281–7,047) [24], slightly larger than the corresponding 4,932 reported COVID-19 deaths [35].

To illustrate the significant differences between excess deaths and reported COVID-19 deaths in various jurisdictions, we plot the excess deaths against confirmed deaths for various countries and US states as of December 10, 2020 in Fig. 3. We observe in Fig. 3(a) that the number of excess deaths in countries like Mexico, Russia, Spain, Peru, and Ecuador is significantly larger than the corresponding number of confirmed COVID-19 deaths. In particular, in Russia, Ecuador, and Spain the number of excess deaths is about three times larger than the number of reported COVID-19 deaths. As described in the Methods section, for certain countries (*e*.*g*., Brazil) excess death data is not available for all states [27]. For the majority of US states the number of excess deaths is also larger than the number of reported COVID-19 deaths, as shown in Fig. 3(b). We performed a least-square fit to calculate the proportionality factor *m* arising in 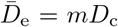 and found *m* ≈ 1.132 (95% CI 1.096– 1.168). That is, across all US states, the number of excess deaths is about 13% larger than the number of confirmed COVID-19 deaths.

**Fig. 3:**
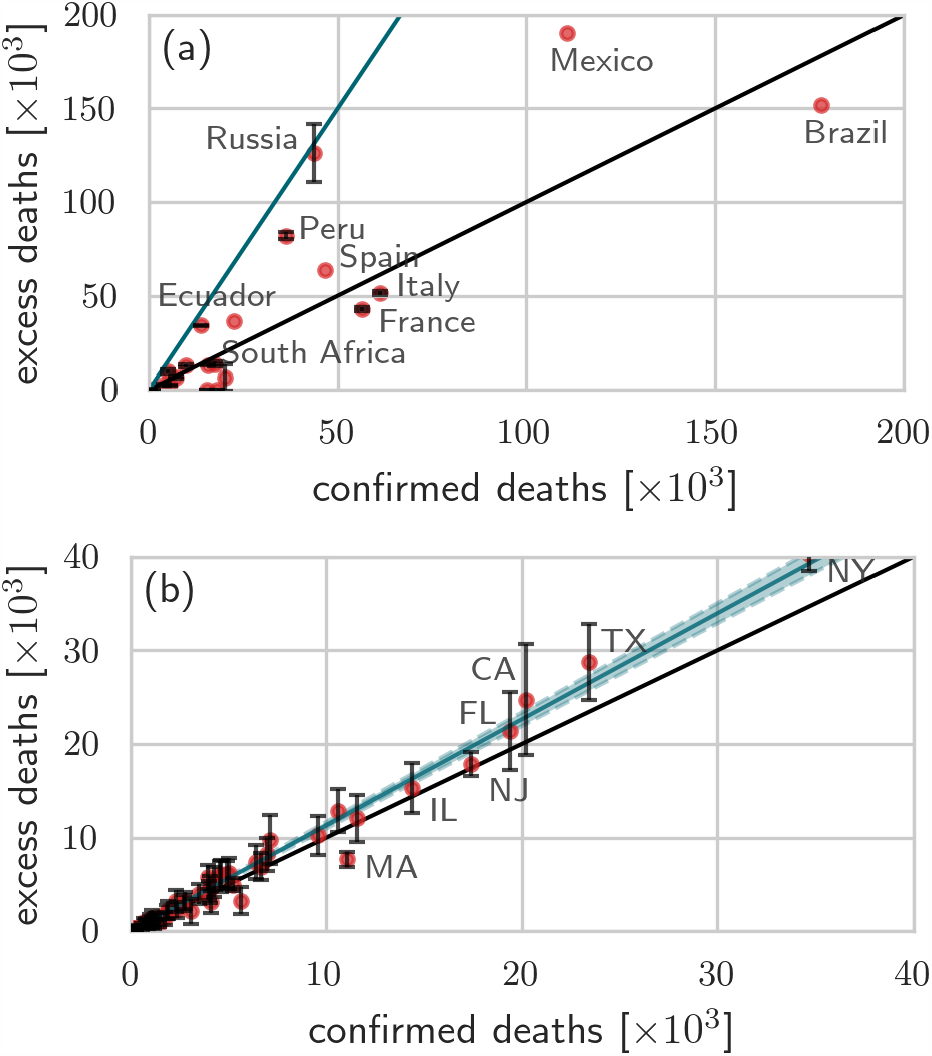
Excess deaths versus confirmed deaths across different countries/states. The number of excess deaths in 2020 versus confirmed deaths across different countries (a) and US states (b). The black solid lines in both panels have slope 1. In (a) the blue solid line is a guide-line with slope 3; in (b) the blue solid line is a least-squares fit of the data with slope 1.132 (95% CI 1.096–1.168; blue shaded region). All data were updated on December 10, 2020 [20, 27, 29, 36].

### Estimation of mortality measures and their uncertainties

We now use excess death data and the statistical and modeling procedures to estimate mortality measures *Z* = IFR, CFR, *M*, ℳ across different jurisdictions, including all US states and more than two dozen countries.^1^. Accurate estimates of the confirmed *N*_c_ and dead *D*_c_ infected are needed to evaluate the CFR. Values for the parameters *Q*, FPR, FNR, and *b* are needed to estimate *N*_c_+*N*_u_ = *f N* in the denominator of the IFR, while 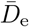 is needed to estimate the number of infection-caused deaths *D*_c_ + *D*_u_ that appear in the numerator of the IFR and ℳ. Finally, since we evaluate the resolved mortality, through Eq. 8, estimates of 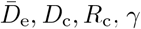, and FPR, FNR (to correct for testing inaccuracies in *D*_c_ and *R*_c_) are necessary. Whenever uncertainties are available or inferable from data, we also include them in our analyses.

Estimates of excess deaths and infected populations themselves suffer from uncertainty encoded in the variances 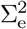 and 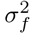. These uncertainties depend on uncertainties arising from finite sampling sizes, uncertainty in bias *b* and uncertainty in test sensitivity and specificity, which are denoted 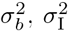, and 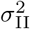, respectively. We use Σ^2^ to denote population variances and *σ*^2^ to denote parameter variances; covariances with respect to any two variables *X, Y* are denoted as Σ_*X,Y*_. Variances in the confirmed populations are denoted 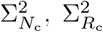, and 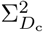 also depend on uncertainties in testing parameters 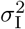 and 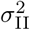. The most general approach would be to define a probability distribution or likelihood for observing some value of the mortality index in [*Z, Z* + d*Z*]. As outlined in the SI, these probabilities can depend on the mean and variances of the components of the mortalities, which in turn may depend on hyperparameters that determine these means and variances. Here, we simply assume uncertainties that are propagated to the mortality indices through variances in the model parameters and hyperparameters [37]. The squared coefficients of variation of the mortalities are found by linearizing them about the mean values of the underlying components and are listed in Table II.

To illustrate the influence of different biases *b* on the IFR we use 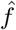 from Eq. (6) in the corrected 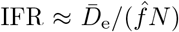. We model RT-PCR-certified COVID-19 deaths [38] by setting the FPR = 0.05 [39] and the FNR = 0.2 [40, 41]. The observed, possibly biased, fraction of positive tests 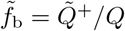 can be directly obtained from corresponding empirical data. As of November 1, 2020, the average of 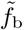 over all tests and across all US states is about 9.3% [42]. The corresponding number of excess deaths is 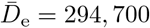 [27] and the US population is about *N* ≈ 330 million [43]. To study the influence of variations in 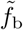, in addition to 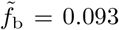, we also use a slightly larger 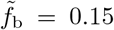 in our analysis. In Fig. 4 we show the apparent and corrected IFRs for two values of 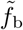 [Fig. 4(a)] and the coefficient of variation CV_IFR_ [Fig. 4(b)] as a function of the bias *b* and as made explicit in Table I. For unbiased testing [*b* = 0 in Fig. 4(a)], the corrected IFR in the US is 1.9% assuming 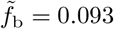 and 0.8% assuming 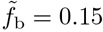. If *b >* 0, there is a testing bias towards the infected population, hence, the apparent 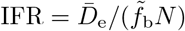 is smaller than the corrected IFR as can be seen by comparing the solid (corrected IFR) and the dashed (apparent IFR) lines in Fig. 4(a). For testing biased towards the uninfected population (*b <* 0), the corrected IFR may be smaller than the apparent IFR. To illustrate how uncertainty in FPR, FNR, and *b* affect uncertainty in IFR, we evaluate CV_IFR_ as given in Table II.

**Fig. 4:**
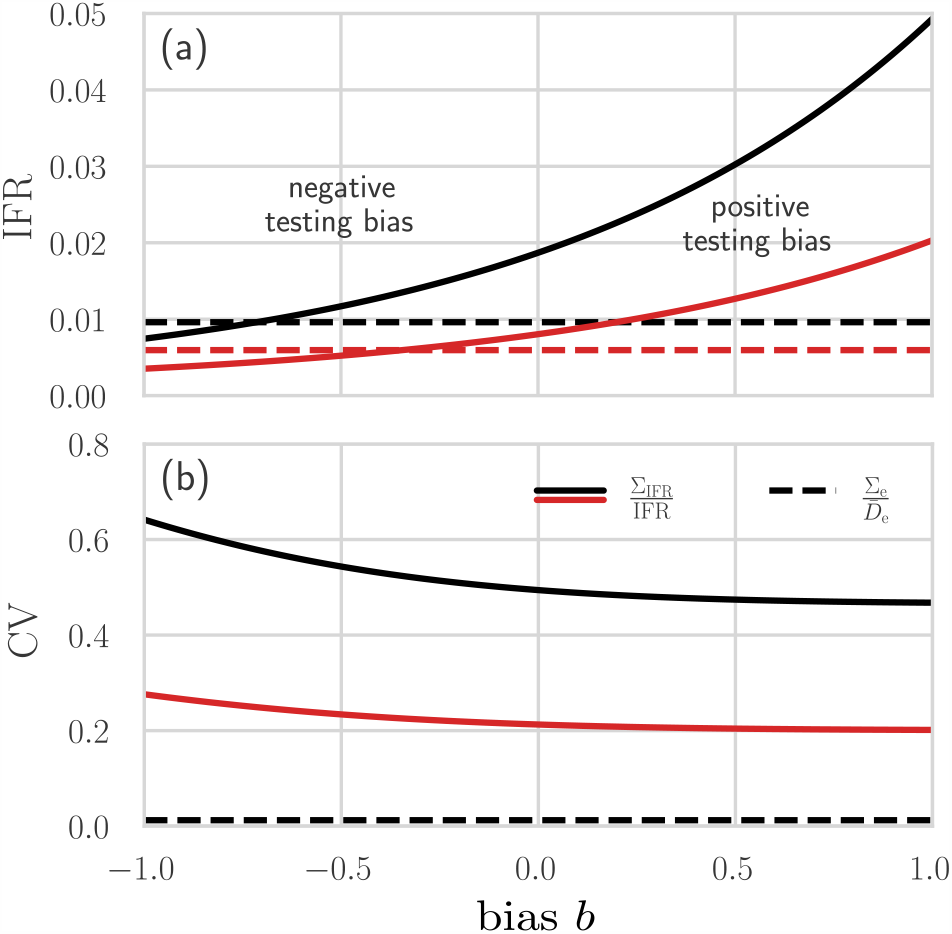
Different mortality measures across different regions. (a) The apparent (dashed lines) and corrected (solid lines) IFR in the US, as of November 1, 2020, estimated using excess mortality data. We set 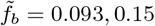, 0.15 (black,red), FPR = 0.05, FNR = 0.2, and *N* = 330 million. For the corrected IFR, we use 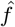 as defined in Eq. (6). Unbiased testing corresponds to setting *b* = 0. For *b >* 0 (positive testing bias), infected individuals are overrepresented in the sample population. Hence, the corrected IFR is larger than the apparent IFR. If *b* is sufficiently small (negative testing bias), the corrected IFR may be smaller than the apparent IFR. (b) The coefficient of variation of *D*_e_ (dashed line) and IFR (solid lines) with *σ*_I_ = 0.02, *σ*_II_ = 0.05, and *σ*_*b*_ = 0.2 (see Tab. II).

The first term in uncertainty 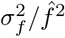 given in Eq. (A6) is proportional to 1*/Q* and can be assumed to be negligibly small, given the large number *Q* of tests administered. The other terms in Eq. (A6) are evaluated by assuming *σ*_*b*_ = 0.2, *σ*_I_ = 0.02, and *σ*_II_ = 0.05 and by keeping FPR = 0.05 and FNR = 0.2. Finally, we infer Σ_e_ from empirical data, neglect correlations between *D*_e_ and *N*, and assume that the variation in *N* is negligible so that Σ_e,*N*_ = Σ_*N*_ ≈ 0. Fig. 4(b) plots CV_IFR_ and 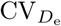 in the US as a function of the underlying bias *b*. The coefficient of variation 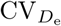 is about 1%, much smaller than CV_IFR_, and independent of *b*. For the values of *b* shown in Fig. 4(b), CV_IFR_ is between 47–64% for 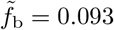 and between 20–27% for 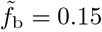.

Next, we compared the mortality measures *Z* =IFR, CFR, *M*, ℳand the relative excess deaths *r* listed in Tab. I across numerous jurisdictions. To determine the CFR, we use the COVID-19 data of Refs. [20, 36]. For the apparent IFR, we use the representation 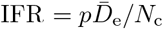 discussed above. Although *p* may depend on the stage of the pandemic, typical estimates range from 4% [44] to 10% [31]. We set *p* = 0.1 over the lifetime of the pandemic. We can also use the apparent 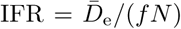, however estimating the corrected IFR requires evaluating the bias *b*. In Fig. 5(a), we show the values of the relative excess deaths *r*, the CFR, the apparent IFR, the confirmed resolved mortality *M*, and the true resolved mortality ℳ for different (unlabeled) regions. In all cases we set *p* = 0.1, *γ* = 100. As illustrated in Fig. 5(b), some mortality measures suggest that COVID-induced fatalities are lower in certain countries compared to others, whereas other measures indicate the opposite. For example, the total resolved mortality ℳfor Brazil is larger than for Russia and Mexico, most likely due to the relatively low number of reported excess deaths as can be seen from Fig. 3 (a). On the other hand, Brazil’s values of CFR, IFR, and *M* are substantially smaller than those of Mexico [see Fig. 5(b)].

**Fig. 5:**
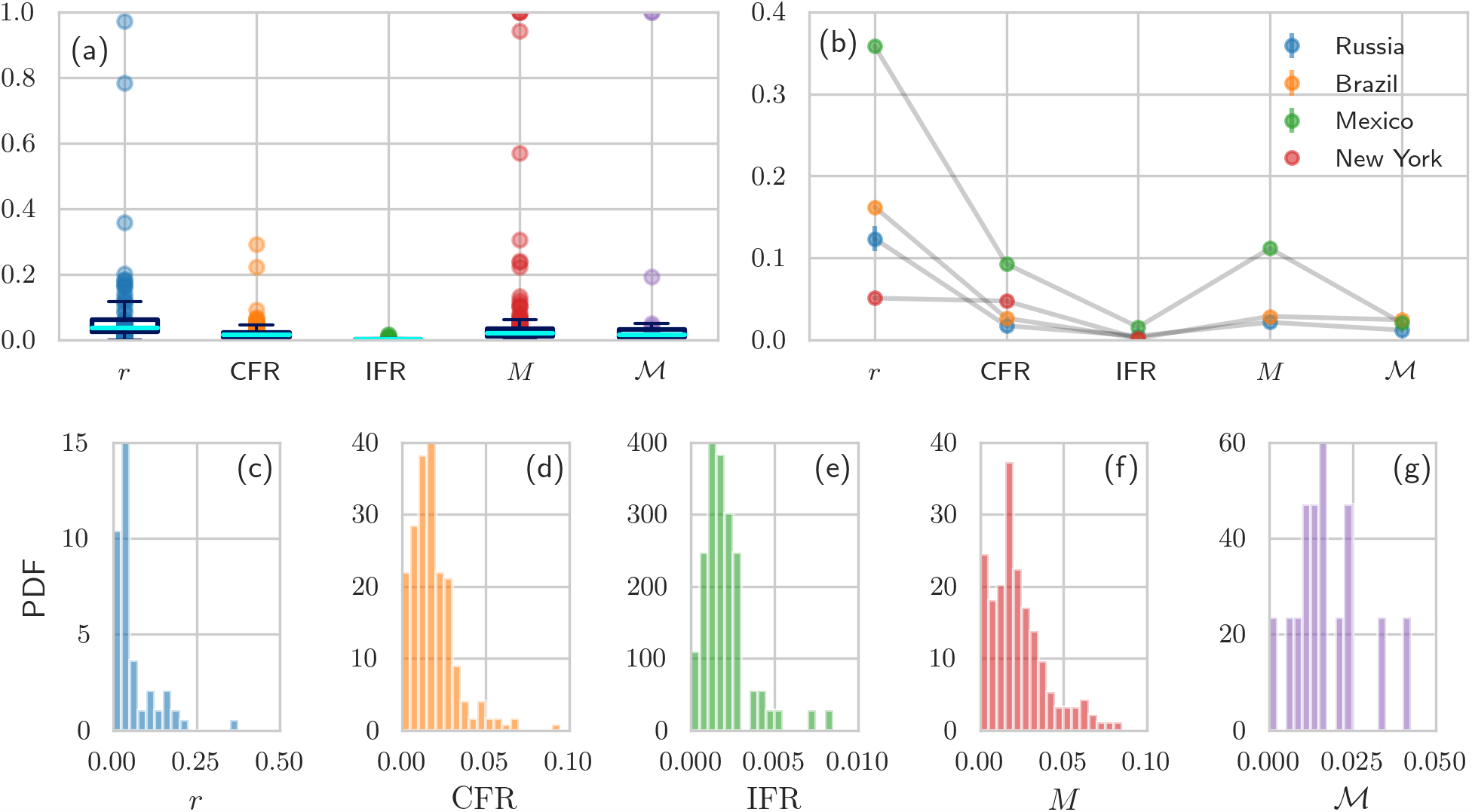
Mortality characteristics in different countries and states. (a) The values of relative excess deaths *r*, the CFR, the 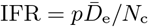 with *p* = *N*_c_*/*(*N*_c_ + *N*_u_) = 0.1 [31], the confirmed resolved mortality *M*, and the true resolved mortality (using *γ* = 100) are plotted for various jurisdictions. (b) Different mortality measures provide ambiguous characterizations of disease severeness. (c–g) The probability density functions (PDFs) of the mortality measures shown in (a) and (b). Note that there are only very incomplete recovery data available for certain countries (*e*.*g*., US and UK). For countries without recovery data, we could not determine *M* and ℳ. The number of jurisdictions that we used in (a) and (c–g) are 77, 246, 73, 191, and 21 for the respective mortality measures (from left to right). All data were updated December 10, 2020 [20, 27, 29, 36].

The distributions of all measures *Z* and relative excess deaths *r* across jurisdictions are shown Fig. 5(c–g) and encode the global uncertainty of these indices. We also calculate the corresponding mean values across jurisdictions, and use the empirical cumulative distribution functions to determine confidence intervals. The mean values across all jurisdictions are 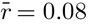 (95% CI 0.0025–0.7800), 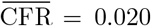 (95% CI 0.0000–0.0565), 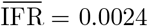 (95% CI 0.0000–0.0150), 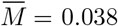 (95% CI 0.0000–0.236), and 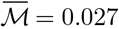 (95% CI 0.000–0.193). For calculating 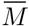 and 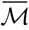, we excluded countries with incomplete recovery data. The distributions plotted in Fig. 5(c–g) can be used to inform our analyses of uncertainty or heterogeneity as summarized in Tab. II. For example, the overall variance 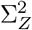 can be determined by fitting the corresponding empirical *Z* distribution shown in Fig. 5(c–g). Table II displays how the related 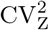 can be decomposed into separate terms, each arising from the variances associated to the components in the definition of *Z*. For concreteness, from Fig. 5(e) we obtain 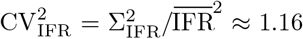 which allows us to place an upper bound on 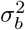 using Eq. (A6), the results of Tab. II, and

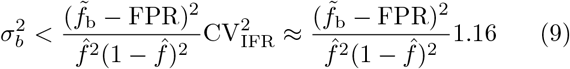

or on 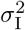 using 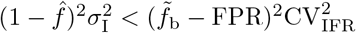.

Finally, to provide more insight into the correlations between different mortality measures, we plot *M* against CFR and ℳ against IFR in Fig. 6. For most regions, we observe similar values of *M* and CFR in Fig. 6(a). Althouigh we expect *M* →CFR and ℳ →IFR towards the end of an epidemic, in some regions such as the UK, Sweden, Netherlands, and Serbia, *M* ≫ CFR due to unreported or incomplete reporting of recovered cases. About 50% of the regions that we show in Fig. 6(b) have an IFR that is approximately equal to ℳ. Again, for regions such as Sweden and the Netherlands, ℳ is substantially larger than IFR because of incomplete reporting of recovered cases.

**Fig. 6:**
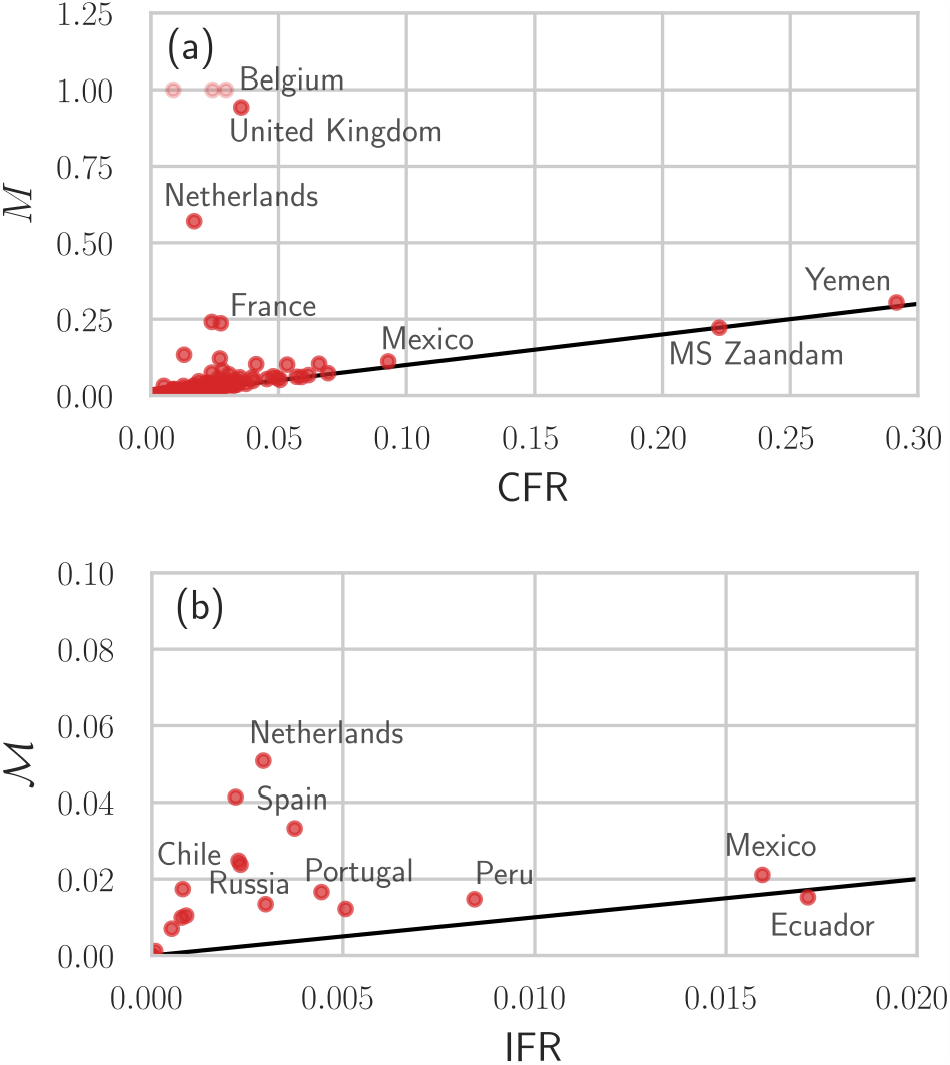
Different mortality measures across different regions. We show the values of *M* and CFR (a) and ℳ (using *γ* = 100) and 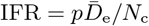 with *p* = *N*_c_*/*(*N*_c_ +*N*_u_) = 0.1 [31] (b) for different regions. The black solid lines have slope 1. If jurisdictions do not report the number of recovered individuals, *R*_c_ = 0 and *M* = 1 [light red disks in (a)]. In jurisdictions for which the data indicate 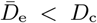, we set 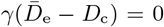 n the denominator of ℳ which prevents it from becoming negative as long as 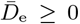. All data were updated on December 10, 2020 [20, 27, 29, 36].

## Discussion

### Relevance

In the first few weeks of the initial COVID-19 outbreak in March and April 2020 in the US, the reported death numbers captured only about two thirds of the total excess deaths [15]. This mismatch may have arisen from reporting delays, attribution of COVID-19 related deaths to other respiratory illnesses, and secondary pandemic mortality resulting from delays in necessary treatment and reduced access to health care [15]. We also observe that the number of excess deaths in the Fall months of 2020 have been significantly higher than the corresponding reported COVID-19 deaths in many US states and countries. The weekly numbers of deaths in regions with a high COVID-19 prevalence were up to 8 times higher than in previous years. Among the countries that were analyzed in this study, the five countries with the largest numbers of excess deaths since the beginning of the COVID-19 outbreak (all numbers per 100,000) are Peru (256), Ecuador (199), Mexico (151), Spain (136), and Belgium (120). The five countries with the lowest numbers of excess deaths since the beginning of the COVID-19 outbreak are Denmark (2), Norway (6), Germany (8), Austria (31), and Switzerland (33) [27] ^2^. If one includes the months before the outbreak, the numbers of excess deaths per 100,000 in 2020 in Germany, Denmark, and Norway are -3209, -707, and -34, respectively. In the early stages of the COVID-19 pandemic, testing capabilities were often insufficient to resolve rapidly-increasing case and death numbers. This is still the case in some parts of the world, in particular in many developing countries [45]. Standard mortality measures such as the IFR and CFR thus suffer from a time-lag problem.

### Strengths and limitations

The proposed use of excess deaths in standard mortality measures may provide more accurate estimates of infection-caused deaths, while errors in the estimates of the fraction of infected individuals in a population from testing can be corrected by estimating the testing bias and testing specificity and sensitivity. One could sharpen estimates of the true COVID-19 deaths by systematically analyzing the statistics of deaths from all reported causes using a standard protocol such as ICD-10 [46]. For example, the mean traffic deaths per month in Spain between 2011-2016 is about 174 persons [47], so any pandemic-related changes to traffic volumes would have little impact considering the much larger number of COVID-19 deaths.

Different mortality measures are sensitive to different sources of uncertainty. Under the assumption that all excess deaths are caused by a given infectious disease (*e*.*g*., COVID-19), the underlying error in the determined number of excess deaths can be estimated using historical death statistics from the same jurisdiction. Uncertainties in mortality measures can also be decomposed into the uncertainties of their component quantities, including the positive-tested fraction *f* that depend on uncertainties in the testing parameters.

As for all epidemic forecasting and surveillance, our methodology depends on the quality of excess death and COVID-19 case data and knowledge of testing parameters. For many countries, the lack of binding international reporting guidelines, testing limitations, and possible data tampering [48] complicates the application of our framework. A striking example of variability is the large discrepancy between excess deaths *D*_e_ and confirmed deaths *D*_c_ across many jurisdictions which render mortalities that rely on *D*_c_ suspect. More research is necessary to disentangle the excess deaths that are directly caused by SARS-CoV-2 infections from those that result from postponed medical treatment [15], increased suicide rates [49], and other indirect factors contributing to an increase in excess mortality. Even if the numbers of excess deaths were accurately reported and known to be caused by a given disease, inferring the corresponding number of unreported cases (*e*.*g*., asymptomatic infections), which appears in the definition of the IFR and ℳ (see Tab. I), is challenging and only possible if additional models and assumptions are introduced.

Another complication may arise if the number of excess deaths is not significantly larger than the historical mean. Then, excess-death-based mortality estimates suffer from large uncertainty/variability and may be meaningless. While we have considered only the average or last values of 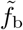, our framework can be straightforwardly extended and dynamically applied across successive time windows, using *e*.*g*., Bayesian or Kalman filtering approaches.

Finally, we have not resolved the excess deaths or mortalities with respect to age or other attributes such as sex, co-morbidities, occupation, etc. We expect that age-structured excess deaths better resolve a jurisdiction’s overall mortality. By expanding our testing and modeling approaches on stratified data, one can also straight-forwardly infer stratified mortality measures *Z*, providing additional informative indices for comparison.

## Conclusions

Based on the data presented in Figs. 5 and 6, we conclude that the mortality measures *r*, CFR, IFR, *M*, and ℳ may provide different characterizations of disease severity in certain jurisdictions due to testing limitations and bias, differences in reporting guidelines, reporting delays, etc. The propagation of uncertainty and coefficients of variation that we summarize in Tab. II can help quantify and compare errors arising in different mortality measures, thus informing our understanding of the actual death toll of COVID-19. Depending on the stage of an outbreak and the currently available disease monitoring data, certain mortality measures are preferable to others. If the number of recovered individuals is being monitored, the resolved mortalities *M* and ℳ should be preferred over CFR and IFR, since the latter suffer from errors associated with the time-lag between infection and resolution [10]. For estimating IFR and ℳ, we propose using excess death data and an epidemic model. In situations in which case numbers cannot be estimated accurately, the relative excess deaths *r* provides a complementary measure to monitor disease severity. Our analyses of different mortality measures reveal that

- The CFR and *M* are defined directly from confirmed deaths *D*_c_ and suffers from variability in its reporting. Moreover, the CFR does not consider resolved cases and is expected to evolve during an epidemic. Although *M* includes resolved cases, its additionally required confirmed recovered cases *R*_c_ add to its variability across jurisdictions. Testing errors affect both *D*_c_ and *R*_c_, but if the FNR and FPR are known, they can be controlled using Eq. (A3) given in the SI.
- The IFR requires knowledge of the true cumulative number of disease-caused deaths as well as the true number of infected individuals (recovered or not) in a population. We show how these can be estimated from excess deaths and testing, respectively. Thus, the IFR will be sensitive to the inferred excess deaths and from the testing (particularly from the bias in the testing). Across all countries analyzed in this study, we found a mean IFR of about 0.24% (95% CI 0.0–1.5%), which is similar to the previously reported values between 0.1 and 1.5% [31–33].
- In order to estimate the resolved true mortality, an additional relationship is required to estimate the unconfirmed recovered population *R*_u_. In this paper, we propose a simple SIR-type model in order to relate *R*_u_ to measured excess and confirmed deaths through the ratio of the recovery rate to the death rate. The variability in reporting *D*_c_ across different jurisdictions generates uncertainty in ℳ and reduces its reliability when compared across jurisdictions.
- The mortality measures that can most reliably be compared across jurisdictions should not depend on reported data which are subject to different protocols, errors, and manipulation/intentional omission. Thus, the per capital excess deaths and relative excess deaths *r* (see last column of Table I) are the measures that provide the most consistent comparisons of disease mortality across jurisdictions (provided total deaths are accurately tabulated). However, they are the least informative in terms of disease severity and individual risk, for which *M* and ℳ are better.
- Uncertainty in all mortalities *Z* can be decomposed into the uncertainties in component quantities such as the excess death or testing bias. We can use global data to estimate the means and variances in *Z*, allowing us to put bounds on the variances of the component quantities and/or parameters.

Parts of our framework can be readily integrated into or combined with mortality surveillance platforms such as the European Mortality Monitor (EURO MOMO) project [28] and the Mortality Surveillance System of the National Center for Health Statistics [21] to assess disease burden in terms of different mortality measures and their associated uncertainty.

## Data Availability

All datasets used in this study are available from
Refs. [21-25]. The source codes used in our analyses are publicly available at [17].

## Data availability

All datasets used in this study are available from Refs. [21–25]. The source codes used in our analyses are publicly available at [17].

## Acknowledgements

LB acknowledges financial support from the Swiss National Fund (P2EZP2 191888). The authors also acknowledge financial support from the Army Research Office (W911NF-18-1-0345), the NIH (R01HL146552), and the National Science Foundation (DMS-1814364, DMS-1814090).

## Supplementary Information

### Examples of excess death data

**Fig. A1:**
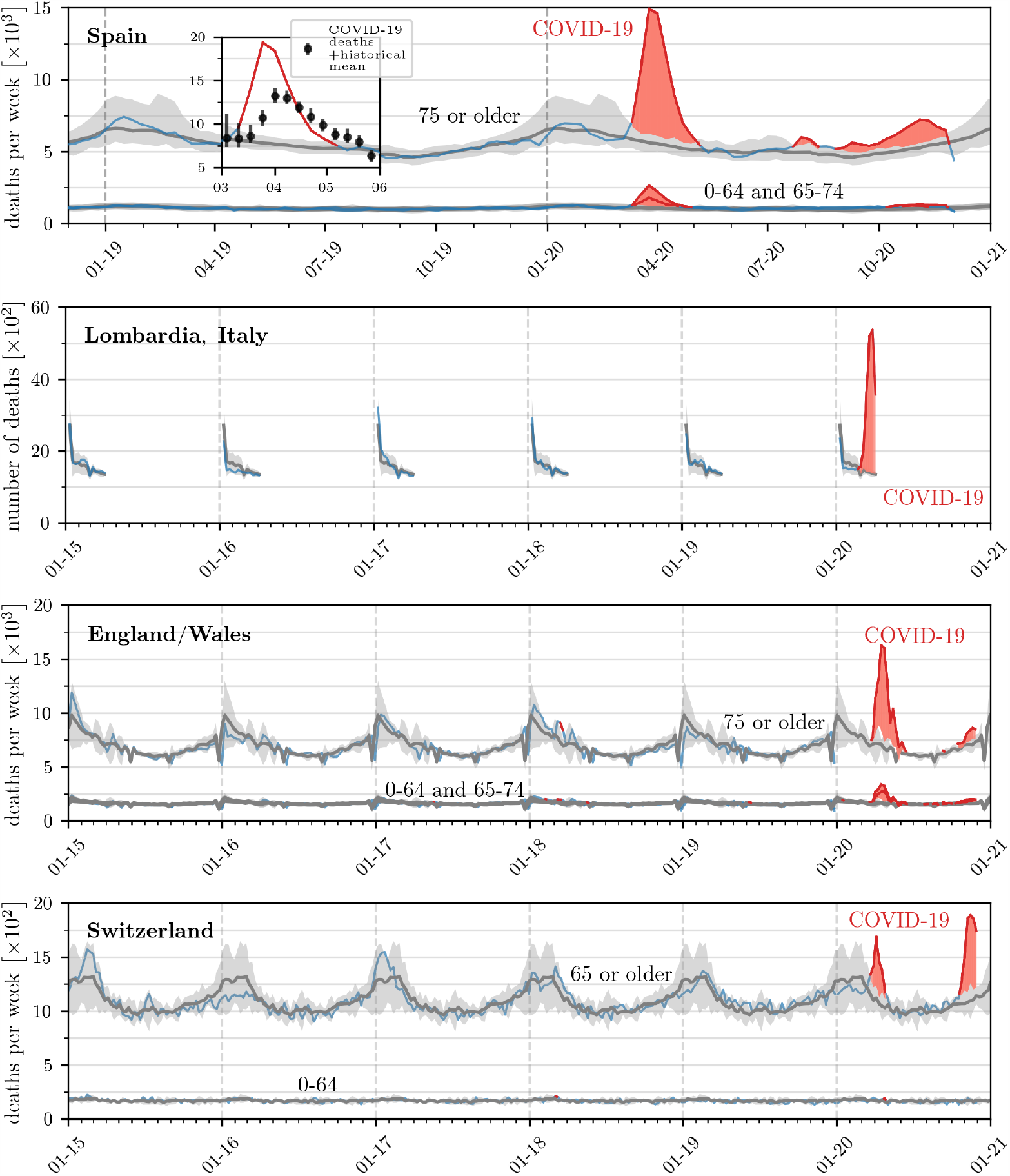
Mortality evolution in different countries. The evolution of weekly deaths in New York City, Spain, England/Wales, and Switzerland for different age classes (where available). Grey solid lines and shaded regions represent the historical mean numbers of deaths and corresponding confidence intervals. Blue solid lines indicate weekly deaths and weekly deaths that lie outside the confidence intervals are indicated by solid red lines. For England/Wales and Switzerland, weekly means and 95% confidence intervals are based on data from 2015–2019. In the case of Spain, we show the reported COVID-19 deaths across all age classes [35] in the inset and use the 99% confidence intervals that are directly provided in the corresponding data [26]. The red shaded regions represent the mean cumulative excess deaths *D*_e_. The data are derived from Refs. [21–25].

We tally weekly deaths according to Eq. (1) for each week *i* starting from the first week of 2020, and cumulative excess deaths as in Eq. (2) adding all weekly contributions from the first week of 2020 onwards. Note that some governmental agencies tabulate weekly deaths starting on the Sunday closest to January 1 2020 (December 29 2019, such as the United States), others instead use January 1 2020 as the first day of the week (such as Germany). A detailed list of how each country bins weekly deaths is included in Ref. [27]. The final week *k* up to which the cumulative count is taken depends on data availability, since some countries have larger reporting delays than others. In the majority of cases *k* is beyond the fourth week of November 2020. Quantities are calculated from data that include deaths from typically *J* = 5 previous years [27]. In Fig. A2 we plot the weekly confirmed deaths 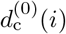, the cumulative deaths 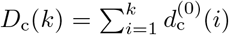, and the mean weekly and cumulative excess deaths 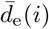 for 2020 as available from data. We also show 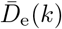 per 100,000 persons from the start of 2020 using Eqs. (1) and (2). The corresponding error bars in Fig. A2 indicate 95% confidence intervals defined by 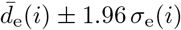 and 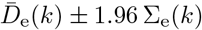 in Eqs. (1) and (2), respectively. For Spain, we used the 99% confidence intervals that are directly provided in the corresponding data [26] to approximate the 95% confidence intervals. Excess death statistics evolve differently across different countries and regions. For example, in France excess deaths were negative until the end of March 2020, quickly increasing in April 2020. In Ecuador and Peru, the number of excess deaths is more than 2.5 times larger than the corresponding number of confirmed COVID-19 deaths.

**Fig. A2:**
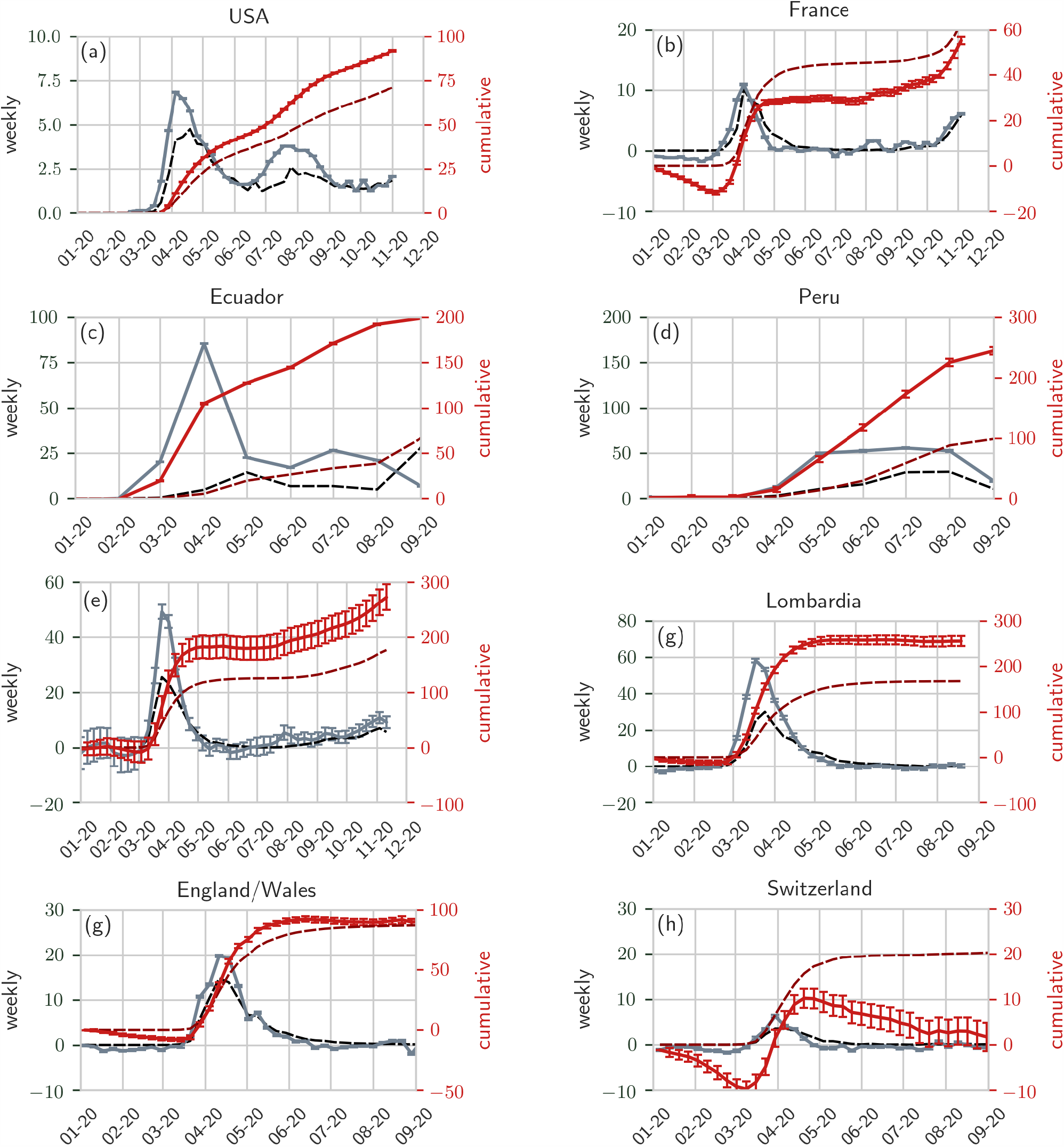
Weekly and cumulative death rates in different countries and regions. We compare the evolution of confirmed weekly deaths 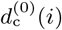 (dashed black curves) and cumulative deaths *D*_c_(*k*) (dashed dark red curves) with weekly excess deaths 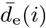 (solid grey curves) and cumulative excess deaths 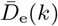 (solid red curves). The deaths are plotted in units of per 100,000 in different countries and regions. The data are derived from Ref. [27] and the error bars for the excess deaths are derived from Eqs. (1) and (2). For Spain, we used the 99% confidence intervals that are directly provided in the corresponding data [26] to approximate the 95% confidence intervals. Typically, we find 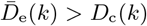.

### Statistical testing model

Given biases in sampling and testing errors, it is important to use a statistical testing model that takes them into account when estimating the fraction *f* of a population *N* that are infected. Testing biases arises, for example, if symptomatic individuals are more likely to seek testing. Thus, the probability *f*_b_ that an individual who chooses to be tested is positive may be different from *f* the probability that a *randomly* selected individual is positive, as defined in Eq. (3). If all tests are error-free, the probability that *Q*^+^ positive results arise from the *Q* ≥ *Q*^+^ administered tests is given by

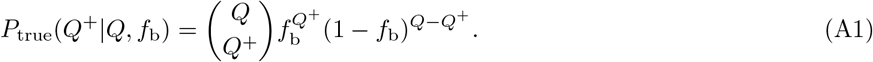

Eq. (A1) is derived under the assumption that once individuals are tested, they are “replaced” in the population and can be tested again. The analogous distribution *P*_true_(*Q*^+^ |*Q, f*_b_) for testing “without replacement” can be straightforwardly derived and yields results quantitatively close to Eq. (A1) provided *Q/N ≲* 0.3.

Eq. (A1) also assumes flawless testing. Tests with Type I (false positives) and Type II (false negatives) may wrongly catalog uninfected individuals as infected (with rate FPR) while missing some infected individuals (with rate FNR). For serological COVID-19 tests, such as antibody tests, the estimated percentages of false positives and false negatives are typically low, with FPR ≈ 0.03 − 0.07 and FNR ≈ 0.1 [39, 50, 51]. For RT-PCR tests, the FNRs depend strongly on the actual assay method [52, 53] and typically lie between 0.1 and 0.3 [40, 41] but might be as high as FNR ≈ 0.68 if throat swabs are used [39, 41]. FNRs can also vary significantly depending on how long after initial infection the test is administered [54]. A systematic review conducted worldwide found FNR ≈ 0.54 at initial testing [55], underlying the need for retesting. Reported percentages of false positives in RT-PCR tests are about FPR ≈ 0.05 [39]. A large meta-analysis of serological tests estimates FPR≈ 0.02 and FNR ≈ 0.02 − 0.16 [54]. These testing errors can lead to inaccurate estimates of disease prevalence; uncertainty in FPR, FNR will thus lead to uncertainty in the estimate of prevalence.

As illustrated through Fig. 2, errors in testing may result in the recorded number 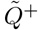 of positive tests to be different from the *Q*^+^ that would be obtained under perfect testing. The probability that 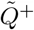 positive tests are returned due to testing errors can be described in terms of *Q*^+^, FPR, and FNR and the corresponding probability distribution 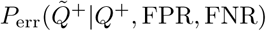 is given by

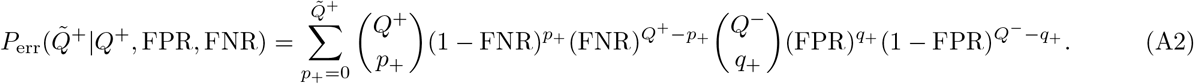

where 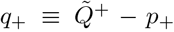. By convolving 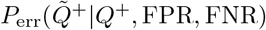 with *P*_true_(*Q*^+^|*Q, f*_b_) we derive the overall likelihood distribution for the measured number 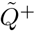 of true *and* false positives given a set of specified parameters *θ* = {*Q, f, b*, FPR, FNR} describing the population and testing

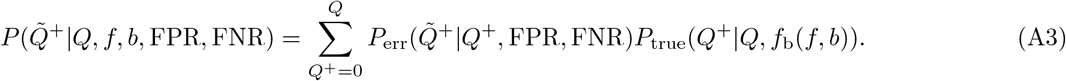

When 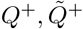, and *Q* ≫ 1, we can approximate *P*_true_, *P*_err_, and *P* by normal distributions and rewrite *P* as a function of the observed positive fraction 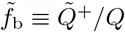 (Eqs. (4) and (5)).

Using Bayes’ rule, we can then formally define the likelihood of *θ* given a measured 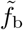,

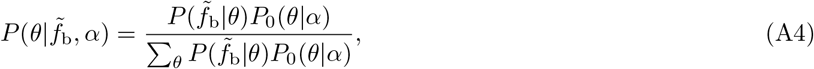

where 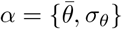 are hyperparameters defining the prior *P*_0_(*θ*|*α*), such as their means 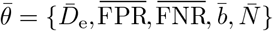and standard deviations *σ*_*θ*_ = {Σ_e_, *σ*_I_, *σ*_II_, *b, σ*_*b*_, *σ*_*N*_}. Formally, the probability of measuring a value of a mortality measure *Z* = CFR, IFR, *M*, ℳ, or *r*, can be computed from

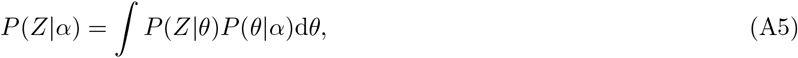

where *P* (*Z* | *θ*) defines the statistical model of the mortality measure given the components and parameters *θ* and the hyperparameters *α* defining the distribution over *θ*. For example, if *Z* is the value of the IFR, *θ* = {*D*_e_, *f, N*} and 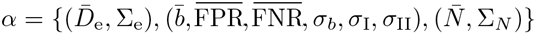 are the mean and standard deviation of excess deaths, testing parameters, and the total population, respectively.

A simpler way to incorporate uncertainty in the infected fraction *f* is to assume a Gaussian approximation for all distributions and propagate the uncertainty in testing parameters. The squared coefficient of variation 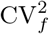 is then decomposed into the parameter variances according to

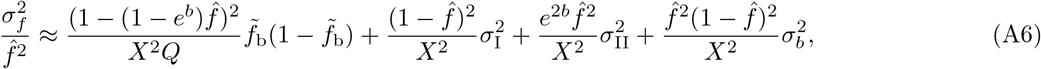

where 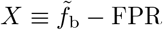. The values of *b*, FPR, FNR above are mean or maximum likelihood estimates of the bias and testing errors, and 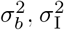 and 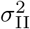 are their associated uncertainties. The means and variances 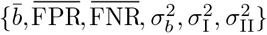 represent hyperparameters associated with testing (see SI). Our result for 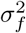 in Eq. (A6) assumes {*b*, FPR, FNR} are uncorrelated. Since *Q* ≫ 1 is typically large, we expect the first contribution to the uncertainty, arising from stochasticity in the sampling and proportional to 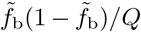 to be negligible. Uncertainties in other quantities will ultimately contribute to uncertainty in the mortalities *Z*, as listed in Table II.

## Modeling of resolved mortality

In Fig. A3, we show the evolution of ℳ for Spain and Lombardia, using different effective recovery rates of unreported cases *γ*. We compute ℳ according to Eq. (8) and use excess mortality data of Fig. A1 to determine 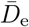. The corresponding data for confirmed recovered and deceased individuals, *R*_c_ and *D*_c_, is taken from Ref. [26]. Current estimates of the IFR are 0.1 − 1.5% [31–33]. To obtain a value of ℳ in a similar range, we vary *γ* from 1 − 1000 and find that ℳ ≈ 0.1 − 1% is consistent with *γ* = 100 − 1000.

**Fig. A3:**
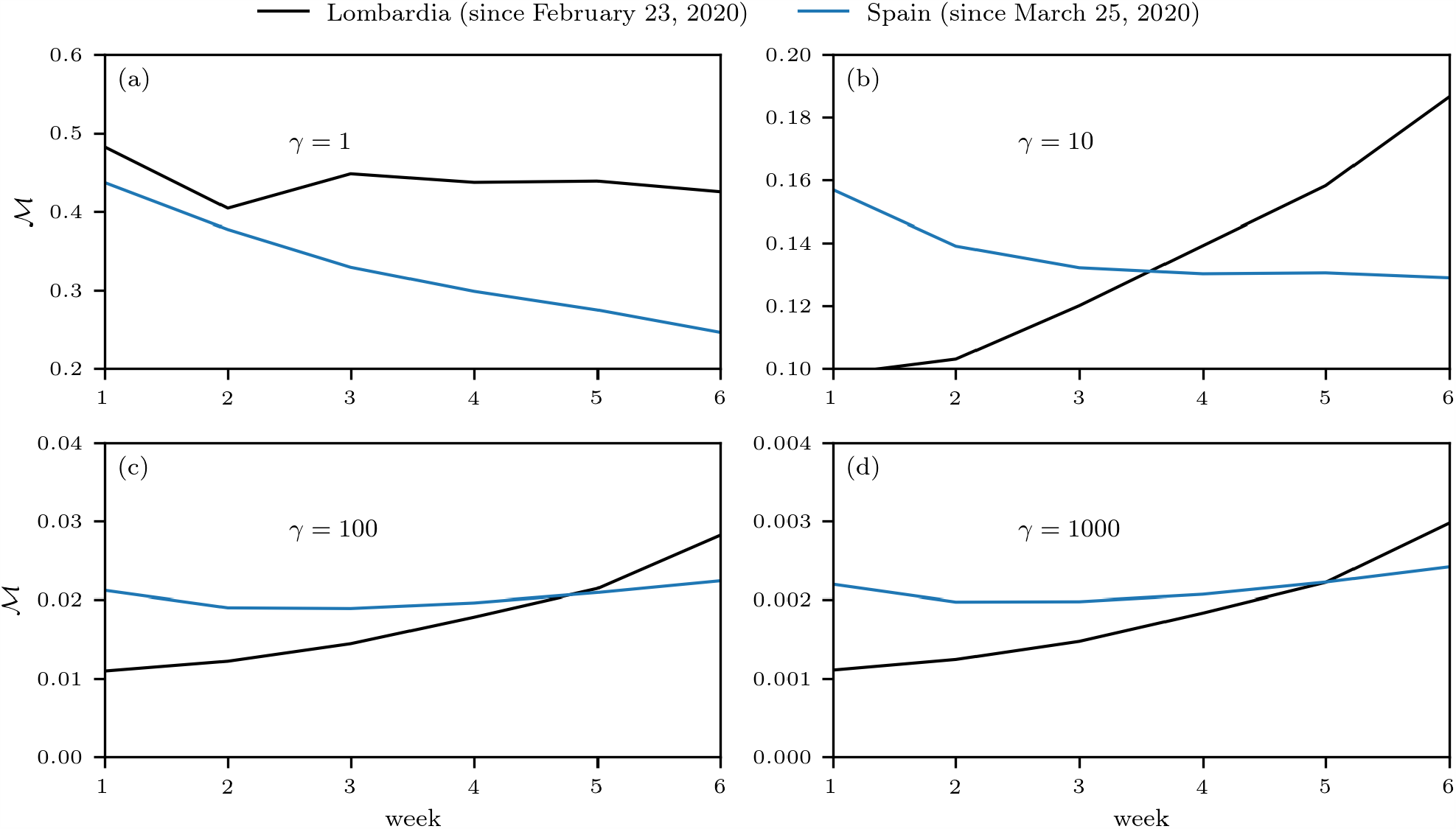
Evolution of resolved mortality. We show the evolution of ℳ (*t*) for different values of effective recovery rates of unreported cases *γ*. The data are derived from Refs. [22, 25].

## Footnotes

1 We provide an online dashboard that shows the real-time evolution of CFR and *M* at https://submit.epidemicdatathon.com/\#/dashboard

2 Note that Switzerland experienced a rapid growth in excess deaths in recent weeks. More recent estimates of the number of excess deaths per 100,000 suggest a value of 64 [26], which is similar to the corresponding excess death value observed in Sweden.

